# Annual rehospitalization rates due to COVID-19 reinfection in Rio de Janeiro: the role of vaccination and associated factors

**DOI:** 10.1101/2025.08.31.25334798

**Authors:** Ranna Kíssia Alves das Neves, Luiz Max Carvalho, Leonardo Soares Bastos, Daniel Antunes Maciel Villela, Valeria Saraceni, Débora Medeiros de Oliveira Cruz, Antonio Guilherme Pacheco

## Abstract

**Introduction:** The COVID-19 pandemic posed significant challenges, including rehospitalization of patients due to SARS-CoV-2 reinfection, particularly in the context of variants of concern and Brazil’s COVID-19 vaccination campaign. This study aimed to estimate the rate of rehospitalizations due to reinfection in the municipality of Rio de Janeiro (2020–2022), assess the impact of individual vaccination status, and identify factors associated with this outcome.

**Methods:** Cohort study including individuals aged 18 years or older who were hospitalized with Severe Acute Respiratory Illness (SARI) resulting from SARS-CoV-2 infection in Rio de Janeiro. Data from four administrative databases (SIPNI, e-SUS Notifica, SIVEP-Gripe, and SIM) were integrated through record linkage. Rehospitalizations due to reinfection were defined as those occurring ≥90 days after the initial hospitalization. Annual rehospitalization rates were calculated per 100 person-years. The analysis included demographics, hospital discharge disposition, and vaccination-related variables. Cox proportional hazards models with deaths considered as competing risks were used.

**Results:** Of the 59,001 individuals included, 1,957 (3.3%) experienced rehospitalization. The main finding was that a complete vaccination regimen with a booster (≥3 doses) was associated with a 39% reduction in the risk of rehospitalization (HR: 0.61; 95% CI: 0.52–0.73), while older age, multimorbidity, and Black race/ethnicity emerged as significant risk factors.

**Conclusion:** Full vaccination with booster doses offered protection against COVID-19-related rehospitalization; however, racial inequities, advanced age, and comorbidities remain important risk factors. Public health policies that expand periodic booster vaccination, strengthen health education, and address social vulnerabilities are urgently needed as a fundamental strategy for preventing future pandemics.

## 1 INTRODUCTION

The COVID-19 pandemic posed a major challenge to the Brazilian healthcare system, characterized by high incidence, widespread transmission, and elevated mortality. It demanded improvements in access to care, quality of services, surveillance, and control strategies^[1]^. In this complex scenario, the possibility of patient rehospitalization emerged as an ongoing concern^[2]^, especially considering the circulation of viral variants of concern (VOCs), which raised questions about vaccine effectiveness and the potential for reinfection^[3]^.

Brazil launched its national COVID-19 vaccination campaign in January 2021, initially with the inactivated virus vaccine CoronaVac and the viral vector vaccine ChAdOx1 nCoV-19/AstraZeneca. Later, the National Immunization Program (PNI) incorporated mRNA vaccines (Pfizer-BioNTech) and viral vector vaccines (Janssen – single dose). While mass vaccination led to a significant reduction in mortality and severe cases, shaping the course of the pandemic—other concurrent factors may have influenced clinical outcomes^[4]^.

In the city of Rio de Janeiro, over 1.2 million confirmed cases, almost 20% of the city’s population, and 38 thousand deaths were recorded by December 2022, alongside more than 18 million vaccine doses administered^[5]^. However, the specific impact of vaccination on reinfection and rehospitalization patterns remain poorly understood.

According to the technical definition of reinfection adopted in Brazil - new symptomatic episodes occurring at least 90 days after the first infection^[6]^ - monitoring

COVID-19 rehospitalizations due to reinfection serves as an important metric to evaluate vaccine-conferred protection and to identify more vulnerable populations. Nonetheless, the literature lacks studies that integrate linked clinical, epidemiological, and vaccination data to investigate this phenomenon, particularly in large urban centers.

Although the World Health Organization has declared the end of the Public Health Emergency of International Concern (PHEIC), it is important to continue monitoring COVID-19 due to the ongoing presence of the virus and its ability to cause severe illness, particularly in unvaccinated individuals and those with comorbidities^[7]^.

In this context, the present study aims to estimate the annual rate of COVID-19 rehospitalizations due to reinfection in the municipality of Rio de Janeiro (2020–2022), assess the impact of individual vaccination status, and identify demographic and clinical factors associated with this outcome.

## 2 MATERIALS AND METHODS

### 2.1 STUDY DESIGN, POPULATION, AND DATA SOURCES

This is a retrospective cohort study based on linked administrative databases, including individuals aged 18 years or older, residing in the municipality of Rio de Janeiro, who were hospitalized and reported with a diagnosis of Severe Acute Respiratory Illness (SARI) due to confirmed or unspecified SARS-CoV-2 infection, in both public and private healthcare sectors, between 2020 and 2022.

The data were accessed in June 2023 and obtained from a database constructed through a previously validated record linkage process. Linkage was performed using an algorithm implemented in Python, comparing name, mother’s name and date of birth among records, as detailed in Pacheco (2008)^[8]^. After the procedure, all databases were anonymized and unique identifiers were attributed to records in the various databases that belong to the same individual. All analyzes were conducted on anonymized datasets.

This database integrated information from four administrative datasets related to the COVID-19 pandemic, provided by the Epidemiological Intelligence Center (CIE) of the municipality of Rio de Janeiro: 1) National Immunization Program Information System (SIPNI) – COVID-19 vaccination; 2) e-SUS Notifica; 3) Influenza Epidemiological Surveillance Information System (SIVEP-Gripe); 4) Mortality Information System (SIM).

To distinguish between rehospitalizations due to reinfection and those resulting from prolonged hospital stays or recurrence of the same infection, a 90-day interval between the date of first symptoms criterion was adopted, following the recommendations of the Brazilian Ministry of Health^[6]^. Thus, only hospitalizations whose symptom ≥ 90 days after the one in the first hospitalization were considered reinfections. This 90-day window was also used to control for immortal time bias, as events such as rehospitalization or death occurring during this period were not counted as time at risk^[9]^. Therefore, follow-up began only 91 days after the first hospitalization (Figure 1).

**Figure 1.**
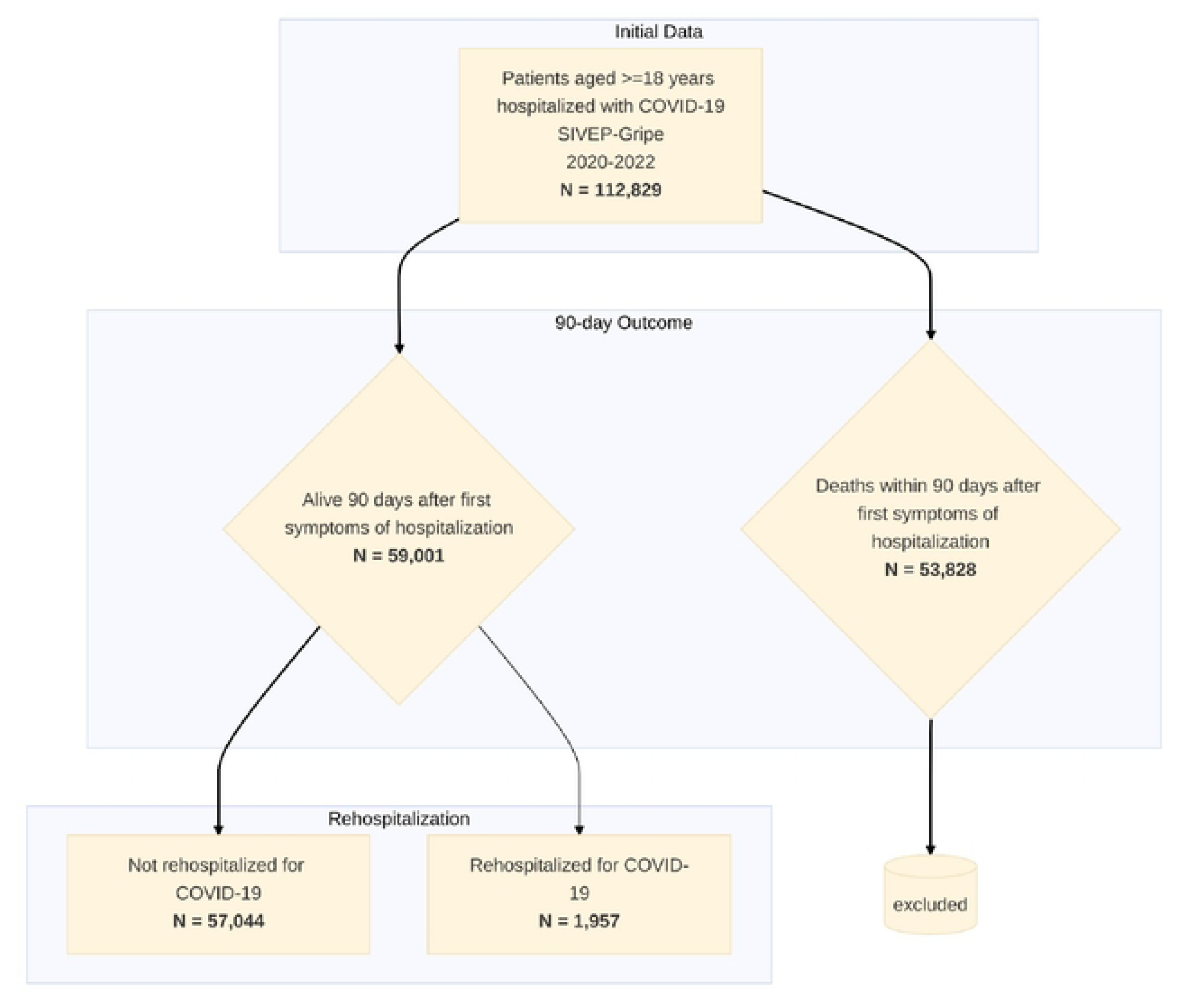
Flowchart of the selection of the cohort of hospitalized COVID-19 patients in Rio de Janeiro (2020–2022)

### 1.1 OUTCOME AND VARIABLES

The primary outcome was the time elapsed between the first hospitalization and the first rehospitalization. The study entry point was defined as the date of symptom onset for the initial hospitalization for COVID-19 or unspecified SARI, as recorded in the SIVEP-Gripe database. Follow-up continued until the first occurrence of: (i) symptoms onset for rehospitalization, (ii) administrative censoring, whichever happened first. Deaths were treated as competing events in the analysis; adjustments were made for potential confounding factors.

The variables used in the analysis included demographic, clinical, vaccination status, and dominant variant data. Demographic variables considered were age (in years), categorized age groups (18–49, 50–69, and ≥ 70 years), sex (male or female), and race/skin color, classified as White; Black (combining Black and Brown individuals); and Others (including Yellow, Indigenous, or missing).

Regarding clinical characteristics, the presence and number of comorbidities (none, one, two, or three or more) were included, based on diseases recorded in the SIVEP-Gripe system. The comorbidities considered encompassed Down syndrome, cardiovascular, pulmonary, renal, hepatic, autoimmune, neurological, hematological diseases, diabetes, and others. Additionally, the final case classification was taken into account (SARI due to COVID-19, unspecified SARI, or missing), admission to the intensive care unit (ICU) — categorized as yes, no, or missing and hospital discharge disposition (cure, death due to COVID-19, death due to other causes, or missing).

Vaccination status was classified into two categories according to the recommendations of the Brazilian Ministry of Health^[10]^: (1) complete vaccination with booster, defined as individuals who received three or more doses of any COVID-19 vaccines, except Janssen, for which two or more doses were considered as a boosted scheme; and (2) other statuses, encompassing unvaccinated individuals, those with incomplete vaccination schedules, or those who completed the primary series without a booster. Vaccination was considered effective starting 14 days after the administration of the last dose received.

An ecological variable was created to determine the dominant SARS-CoV-2 variant. This variable was based on the Epidemiological Bulletins from the municipality of Rio de Janeiro^[5]^, which define the periods of viral variant predominance as follows: (pre-VOC; February 26, 2020 to February 1, 2021), Gamma (February 1, 2021 to June 30, 2021), Delta (June 30, 2021 to November 30, 2021), and Omicron (November 30, 2021 to December 31, 2022).

### 2.3 STATISTICAL ANALYSIS

Descriptive analysis was conducted to compare baseline characteristics between individuals hospitalized once and rehospitalized individuals. Categorical variables were expressed as proportions and compared using Pearson’s chi-square test, while continuous variables were presented as medians and interquartile ranges (IQR) and analyzed using the Kruskal-Wallis test. Rehospitalization rates were calculated annually as the ratio of observed rehospitalizations per year to the total person-years of follow-up, multiplied by 100.

Survival analysis was performed using Cox proportional hazards models with the Fine & Gray competing risks modeling approach^[11]^, which accounts for competing events (such as death) that can preclude the occurrence of the event of interest (rehospitalization). The dominant variant variable was stratified in the model to satisfy the proportional hazards assumption, verified using Schoenfeld residuals. Model selection was based on the Akaike Information Criterion (AIC). This modeling approach allowed for the estimation of hazard ratios (HR) and their respective 95% confidence intervals, adjusted for covariates and time-dependent variables.

All analyses were conducted using the R environment (version 4.2.3)^[12]^, employing the ***survival***^[13,14]^ and ***mstate***^[15,16]^ packages.

### ETHICAL APPROVAL

This study was approved by the Research Ethics Committee of the National School of Public Health Sergio Arouca, Oswaldo Cruz Foundation, Brazil (ENSP/FIOCRUZ; CAAE: 74953523.5.0000.5240), as well as by the Research Ethics Committee of the Municipal Health Department of Rio de Janeiro (SMS/RJ; CAAE: 74953523.5.3001.5279).

## 3 RESULTS

The municipality of Rio de Janeiro reported decreasing COVID-19 readmission rates over time: 5.57 (2020), 2.08 (2021), and 0.72 (2022) per 100 person-years. The analysis comprised a cohort of 59.001 individuals from the original database, selected according to the established inclusion criteria, of whom 1,957 (3.3%) experienced readmission; among those, a higher frequency of females was observed (52%), along with a higher median age (69 vs, 57 years, p < 0.001), a higher proportion of elderly individuals (49% aged 70 or older vs. 33% aged 50–69), and a predominance of black individuals (45% vs. 37%, p < 0.001), as shown in Table 1.

**Table 1.**
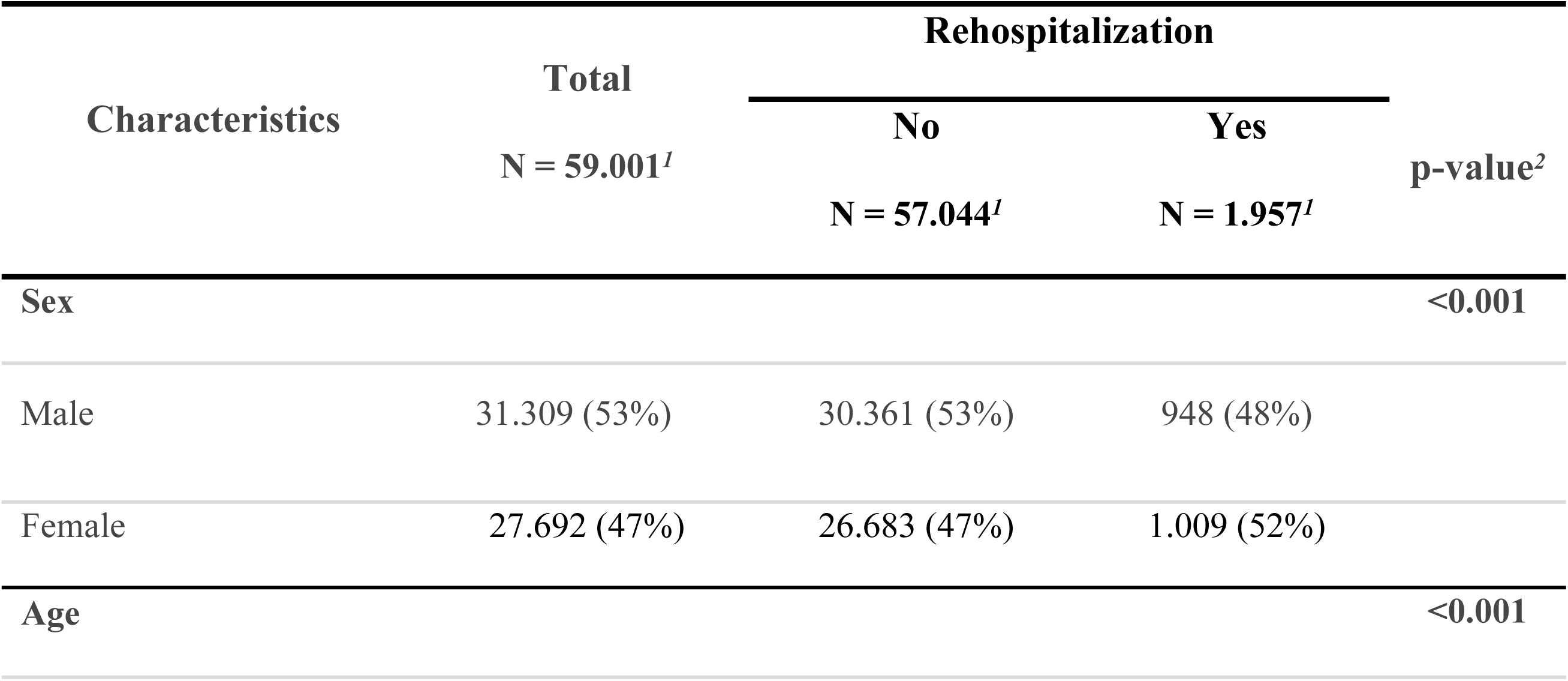
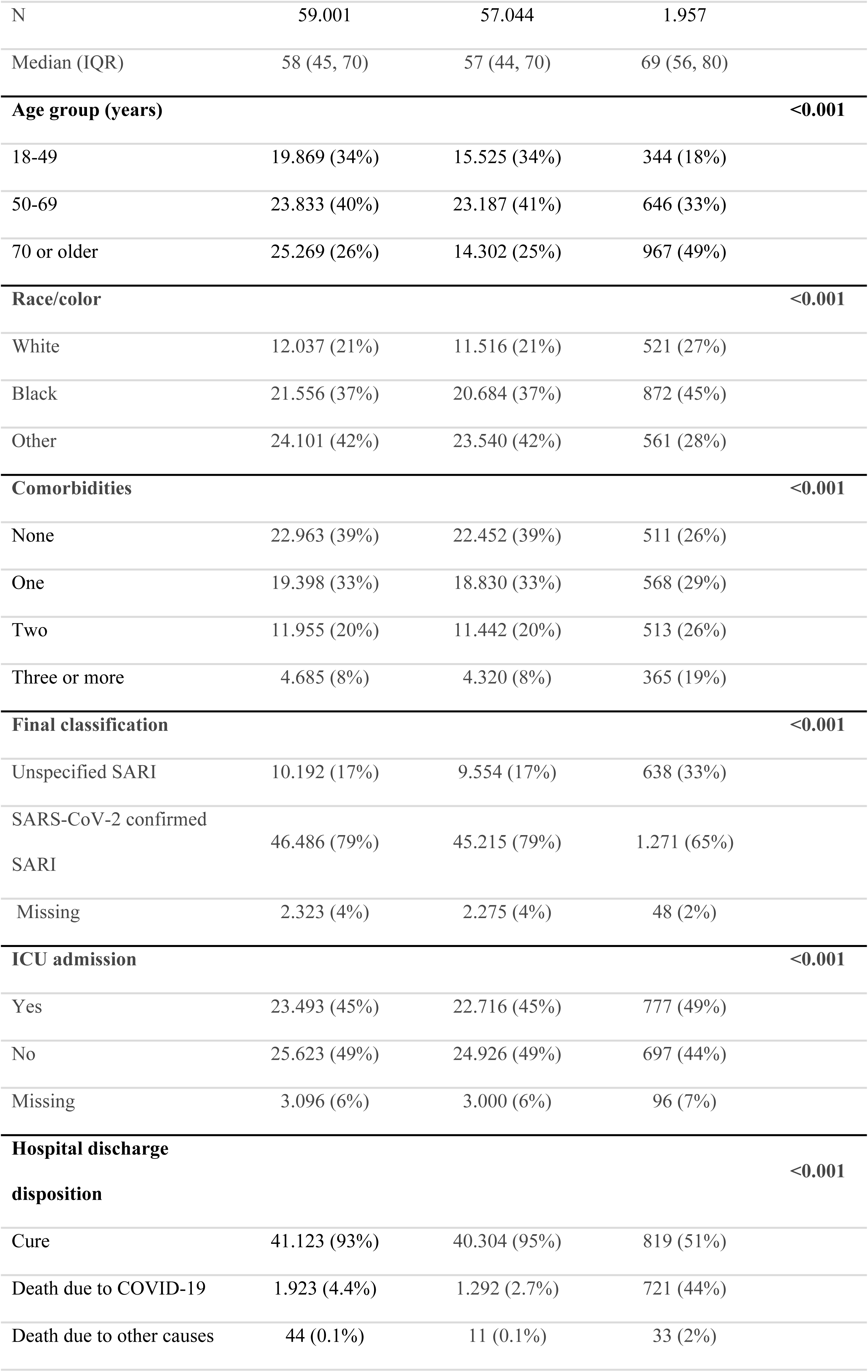
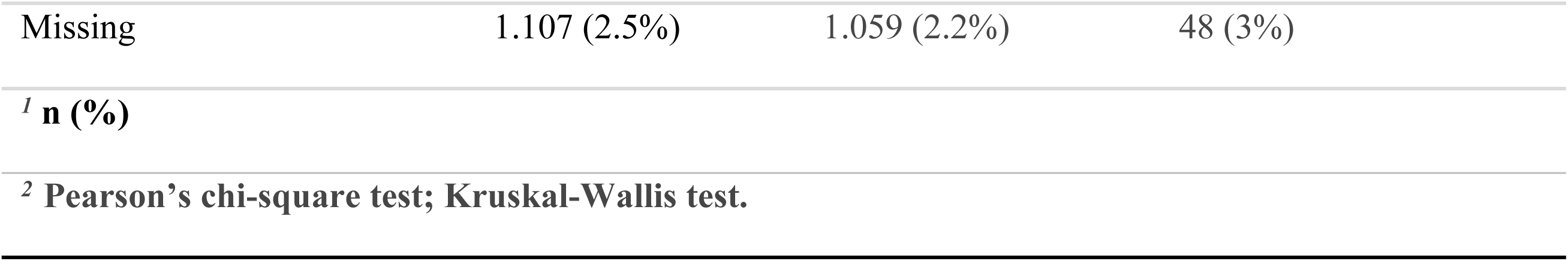
Demographic and clinical characteristics of patients with and without readmission in the municipality of Rio de Janeiro, Brazil, 2020–2022.

Higher prevalence of comorbidities was observed among readmitted patients, with 19% presenting three or more pre-existing conditions, compared to 8% among those not readmitted. The final case classification revealed a greater proportion of unspecified severe acute respiratory illness (SARI) among readmitted patients (33% vs. 17%, p < 0.001). Death was more frequent in this group (44% vs. 2.7%, p < 0.001), as was admission to the intensive care unit (49% vs. 45%, p < 0.001).

Regarding vaccination status, 81% of non-readmitted patients had not received any vaccine prior to their first admission, whereas this proportion was significantly lower among readmitted patients (49%), suggesting a possible protective effect of vaccination against subsequent hospitalization.

Table 2 presents the final model, considering death as a competing event. The main finding was that a complete, boosted vaccination regimen was associated with a significant 39% reduction in the risk of readmission, compared to unvaccinated individuals and those without booster doses.

**Table 2.**
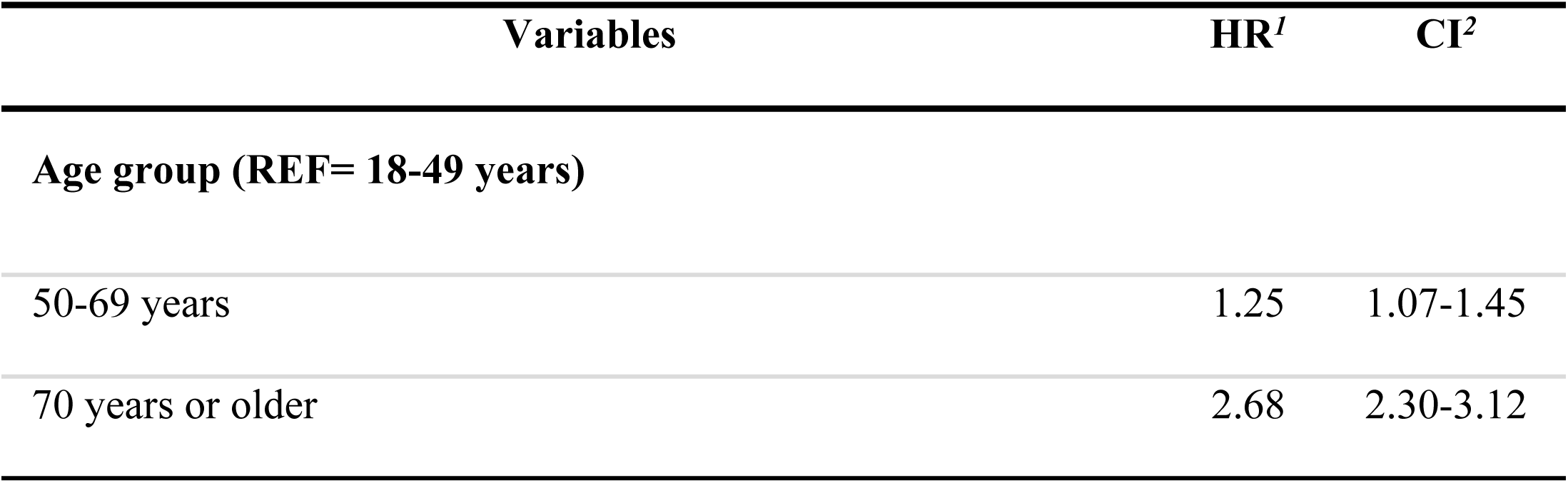
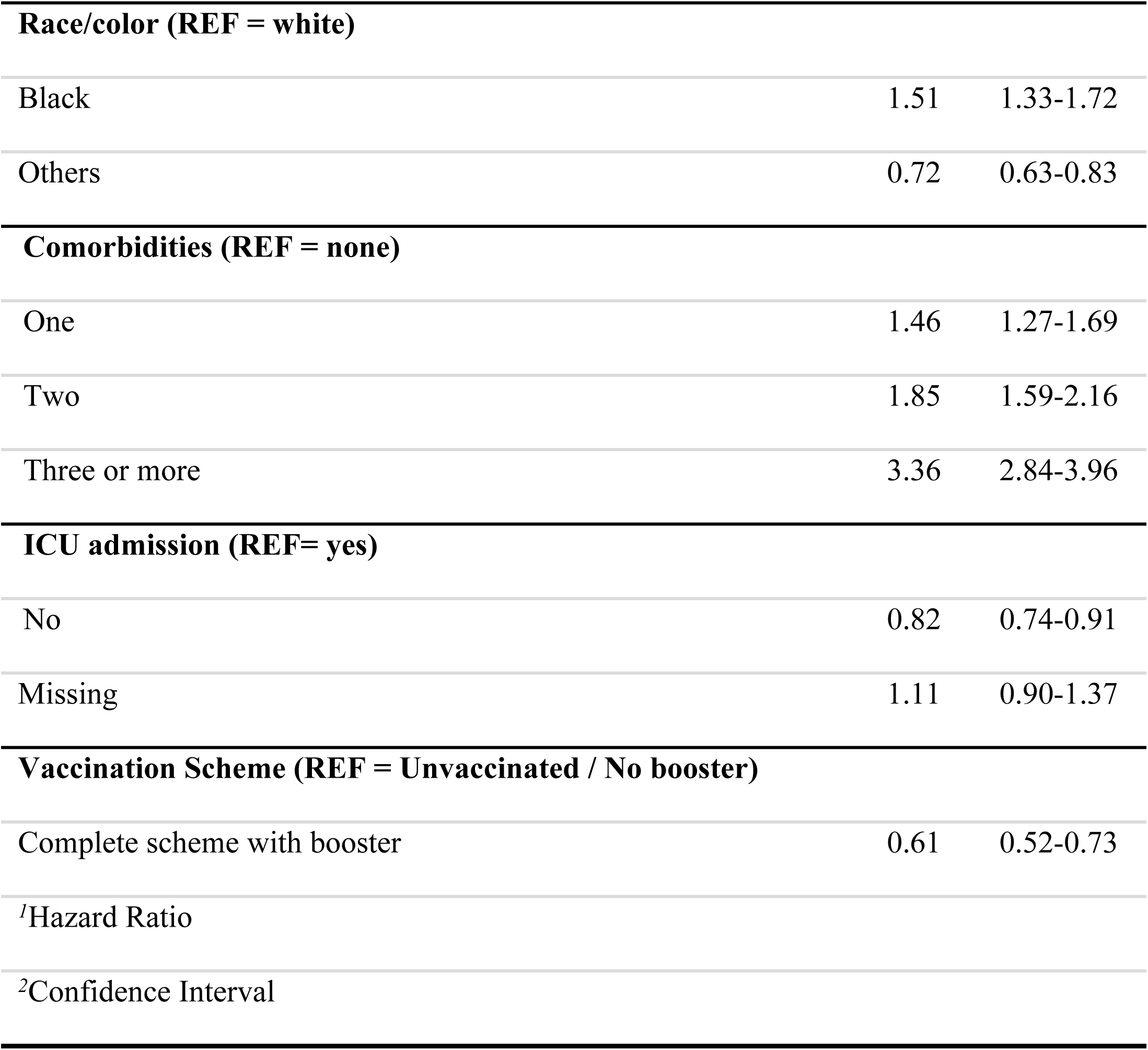
Risk of readmission due to SARS-CoV-2 reinfection, municipality of Rio de Janeiro, 2020–2022.

The dominant variant variable was used to stratify the model to meet the proportional hazards assumption. Although not shown in the table 2, this stratification adjusted the model for the effect of different circulating variants over time, which may influence the risk of readmission.

A progressively increasing risk gradient for readmission was observed according to the number of comorbidities: 46% higher for one, 85% for two, and 236% for three or more, all compared to individuals without comorbidities.

Age was positively associated with risk: individuals aged 50–69 and those 70 years or older had increased risks of 25% and 168%, respectively, compared to the 18– 49 age group.

Regarding race/ethnicity, black individuals showed a 51% higher risk of readmission, while those classified as “other” exhibited a 28% reduced risk, both relative to white individuals. Additionally, not being admitted to the ICU during the first hospitalization was associated with an 18% lower risk of readmission.

## 4 DISCUSSION

This study detailed factors associated with SARS-CoV-2 reinfection-related readmission among hospitalized patients in the municipality of Rio de Janeiro, revealing a landscape marked by social and biological vulnerabilities as well as protective mechanisms. Completion of vaccination schedules, including booster doses, was associated with a 36% reduction in the risk of readmission. Conversely, advanced age, multimorbidity, and Black ethnicity were identified as risk factors. These results underscore the intricate interplay between biological and social determinants in COVID-19 outcomes and reinforce the importance of robust immunization strategies.

Although the COVID-19 literature has predominantly focused on primary outcomes (initial hospitalization or mortality) or readmissions due to worsening of the same infection or post-COVID syndrome, our approach innovates by demonstrating that vaccine protection extends to severe reinfection cases requiring a new hospitalization. This conclusion is supported by national and international evidence: recent Brazilian data show that each additional dose conferred increasing protection against ICU admission, mechanical ventilation, and death, even in elderly individuals with comorbidities^[17]^, while global studies report reductions of 29% and 50% in hospitalization risk with two and three doses, respectively^[18,19]^, with clinical severity predominantly higher among the unvaccinated^[20]^. Additionally, post-infection protection also proved relevant, reducing hospitalizations, readmissions, and post-acute sequelae^[21,22]^, with booster doses further enhancing defense against reinfections^[23–25]^, reinforcing the findings of the present study.

The occurrence of severe outcomes among vaccinated individuals does not contradict the protective effect observed in this study. Rather, it can be partly attributed to waning immunity over time and the circulation of new SARS-CoV-2 variants^[26,27]^. Studies show that booster doses restore vaccine effectiveness^[28–30]^, underscoring their importance in preventing severe cases. Still, coverage remains suboptimal: according to data from the EpiRio Panel (accessed in june 2025), only 63.5% of Rio de Janeiro’s population had received a booster dose^[31]^. This low uptake, combined with vaccine hesitancy^[32]^ and inequalities in access^[33]^, increases the risk of adverse outcomes and facilitates the emergence of new variants and outbreaks.

Advanced age and comorbidities are established risk factors associated with COVID-19 rehospitalization due to reinfection, also demonstrated by Colaneri *et al.* (2025)^[34]^. A progressive increase in risk is observed from the age of 50 onward, as demonstrated in multiple studies^[35–37]^. This pattern can be explained by immunosenescence, the physiological decline in immune response associated with aging^[38,39]^, which impairs viral clearance, increases susceptibility to complications ^[40,41]^, and accelerates the progression of chronic conditions^[42]^.

The simultaneous presence of multiple comorbidities contributes to a pro-inflammatory state and elevated risk of cytokine storm^[43]^, while immunosenescence hinders infection control^[44]^. Our findings corroborate this pattern, revealing a 3.34-fold increased risk of rehospitalization among patients with three or more comorbidities, a result consistent with previous cohort studies^[45–47]^, reinforcing the risk gradient according to the number of pre-existing conditions in relation to rehospitalization.

Concerning race/ethnicity, black race emerged as a risk factor for reinfection-related readmission, consistent with prior data showing black patients having higher risk of readmission^[48,49]^ and higher mortality and hospitalization rates in Brazil^[50,51]^. These disparities reflect the complex intersections between race, socioeconomic status, and healthcare access^[52]^, especially in contexts such as Rio de Janeiro, where 22% of the population live in high social vulnerability areas (favelas)^[53]^. The higher prevalence of comorbidities in readmitted black patients^[54]^ also worsens this outcome. These findings underscore the syndemic nature of COVID-19, where the interplay of social inequities and preexisting vulnerabilities intensifies the clinical and social impact of the disease, exacerbating risks among already marginalized groups. Although racial disparities have been identified, the unexpected protective effect in the “Others” category should be noted, as it includes many cases with missing race data and a few Indigenous and Asian individuals.

No ICU admission during the initial hospital stay was associated with a lower risk of readmission, likely due to milder illness that did not require intensive care. ICU is typically for critical cases, while general wards handle less severe ones^[55,56]^. Additionally, SARS-CoV-2 reinfections are often milder, particularly in individuals fully vaccinated with boosters^[57,58]^.

This study has some limitations. The use of administrative databases may be subject to underreporting and inconsistencies; however, the integration of four complementary data sources enabled cross-validation of the information. The definition of reinfection based on a minimum interval of 90 days does not fully capture the complexity of individual immune responses. Nevertheless, it constitutes a standardized, conservative criterion validated by the Brazilian Ministry of Health, helping to minimize the misclassification of persistent infections as reinfections. Since this definition introduces a bias, immortal time bias, the strategy of initiating follow-up only after this 90-day interval was adopted. This approach was essential to reduce potential biases and enhance the robustness of the results. The ecological classification of viral variants provides an aggregated view of the epidemiological context and may not accurately reflect each patient’s actual exposure to the circulating strain at the time of reinfection. Nevertheless, this approach is widely employed in observational studies, especially in settings where individual-level genomic sequencing data is unavailable. Regarding sample classification, hospitalizations without COVID-19 confirmation (such as unspecified severe acute respiratory syndrome) were included because this classification may have resulted from diagnostic problems and delays or failures in notifications during the pandemic^[59]^. Finally, the absence of direct socioeconomic variables limits a more in-depth analysis of social inequalities. However, this limitation was partially addressed through the inclusion of race/ethnicity, which is recognized as a valid proxy for inequality in the Brazilian context^[60]^.

Despite these limitations, the study offers important methodological strengths. The retrospective cohort design with a large sample size, combined with linkage of four official databases, provided robust statistical power and population representativeness. Additionally, this linkage improved the completeness and accuracy of the information, enabling more reliable analyses. The statistical analysis employed advanced methods, including Fine & Gray competing risks models^[11]^, which appropriately adjusted for death as competing events. Inclusion of individual vaccination variables obtained from the national immunization system enabled real-world vaccine effectiveness evaluation. Furthermore, focusing on reinfection-related readmission fills a critical gap in the literature, which often addresses primary outcomes or readmissions for other causes, providing evidence to guide booster vaccination policies, especially in higher-risk groups.

## 5 CONCLUSION

In summary, this study points out factors associated with SARS-CoV-2 reinfection-related readmission in Rio de Janeiro. The findings reaffirm the role of full vaccination with booster doses in reducing severe outcomes, demonstrating significant protection against readmissions. However, the presence of risk factors such as advanced age, multimorbidity, and racial disparities underscores that vulnerability to COVID-19 is a complex and syndemic phenomenon, intrinsically linked to preexisting social and health determinants.

These findings highlight the urgent need for more equitable public policies that expand access to booster vaccination and promote health education initiatives, particularly targeting the most vulnerable populations. It is essential that such strategies address both clinical risks and social barriers, fostering adherence to preventive measures. Furthermore, given the continuous evolution of the virus and the potential emergence of new variants, periodic revaccination emerges as a crucial measure for COVID-19 control and the mitigation of its future impacts.

## Data Availability

The data cannot be publicly shared due to confidential information contained therein. The data are available from the Municipal Health Secretariat of Rio de Janeiro (contact via the institution’s official email) for researchers who meet the criteria for access to confidential data.

## ACKNOWLEDGMENTS

We thank the Municipal Health Secretariat of Rio de Janeiro and the Municipal Epidemiological Intelligence Center of Rio de Janeiro for providing the dataset used in this study. The authors also acknowledge the support from the National School of Public Health (ENSP/FIOCRUZ), the Scientific Computing Program (PROCC/FIOCRUZ), and the Public Health Epidemiology Program of ENSP for their institutional contributions.

